# The impact of the coronavirus disease 2019 (COVID-19) pandemic on university students’ dietary intake, physical activity, and sedentary behaviour

**DOI:** 10.1101/2021.01.12.21249608

**Authors:** Leandy Bertrand, Keely A. Shaw, Jongbum Ko, Dalton Deprez, Philip D. Chilibeck, Gordon A. Zello

## Abstract

University students are a vulnerable group for poor dietary intake, insufficient physical activity and sedentary behaviour. The purpose of this study was to examine the impact of COVID-19 on university students’ dietary intake, physical activity and sedentary behaviour. Participants were students (n=125) from the Universities of Saskatchewan and Regina. An online questionnaire was administered retrospectively (for pre-pandemic) and prospectively (during the pandemic) to examine students’ dietary intake, physical activity and sedentary behaviour. Overall, nutrient and caloric intakes were significantly reduced (p<0.05) during the pandemic, and alcohol intake increased (p=0.03). Before the pandemic, 16% and 54% of the participants were meeting the Canadian 24-Hour Movement Guidelines for Adults (18–64 years) of 150 minutes of moderate-vigorous physical activity and 8 hours or less of sedentary activity respectively. Only 10% met the guidelines for physical activity while 30% met the guidelines for sedentary behaviour during the pandemic. The minutes per week spent engaging in moderate to vigorous physical activity during the pandemic decreased by approximately 20% (p<0.001). The hours spent in sedentary activities increased by 3 hours per day (p<0.001). Our findings confirm that during the pandemic, students’ inadequate dietary intake, high alcohol consumption, low physical activity and high sedentary behaviour were significantly compounded.

**Novelty:**

- During COVID-19, the nutrient and caloric intakes of university students decreased, and alcohol intake increased significantly.
- University students’ physical activity levels decreased, and sedentary activity increased significantly during COVID-19.
- During COVID-19 students did not engage in sufficient physical activity to offset the increased sedentary behaviour.

## Introduction

The experience of a pandemic may have widespread implications for peoples’ health. In March 2020, the World Health Organization (WHO) declared the rapidly spreading novel coronavirus disease (COVID-19) as a pandemic (WHO, 2020). As the number of confirmed cases continued to increase in Canada, a variety of strict preventative measures were imposed to curb the spread of the virus, including social distancing protocols, quarantines, lockdowns, curfews and closures of non-essential businesses (Government of Canada, 2020). By mid-March 2020, the Government of Saskatchewan declared a provincial State of Emergency in response to the rapidly increasing COVID-19 pandemic (Government of Saskatchewan, 2020). The Universities of Saskatchewan and Regina transitioned to remote teaching while in the wider public grocery stores reduced their hours of operation and capacity for shoppers. Many restaurants (dine-in and takeout) suspended their operations and access to recreational facilities within universities, private establishments and public settings were also shut down as part of the preventative measures imposed (Government of Saskatchewan. 2020). For students within the province, complying with these preventative measures meant experiencing a myriad of changes such as remote learning, home confinement, reduced mobility both by foot and vehicle, halted access to recreation and physical activity facilities, and disruptions in their access to food.

The COVID-19 pandemic has been generating considerable interest in terms of its impact on food intake, sedentary behaviours and level of physical activity. Evidence from a multi-continental survey (n=1047) among adults indicated that during the COVID-19 pandemic daily sitting time increased by 28.6% and frequency and duration of physical activity decreased by 24% and 33.5% respectively (Ammar et al., 2020). The study reported significant increases in consumption of unhealthy food, eating out of control and snacking between meals (Ammar et al., 2020). Specific to the Canadian population, lower physical activity levels and increased sedentary behaviours were reported among a cross-sectional sample of children (n=1472) aged 5-17 years (Moore et al., 2020). Among Canadian adults (n=268), the COVID-19 pandemic resulted in decreased physical activity among inactive (40.5%) and active individuals (22.4%) (Lesser and Neinhuis, 2020).

In Canada, research on university students in a pre-pandemic condition has drawn attention to the reality that they are a vulnerable group for inadequate diet and physical activity. In a recent study at a university in the province of Quebec, 81.2% of students (n=1989) surveyed did not consume the recommended daily servings of fruits and vegetables whilst 55.2% of students were not meeting the Canadian 24-Hour Movement Guidelines for Adults of 150 minutes of moderate to vigorous physical activity per week (Busque et al., 2017). In an earlier study from another Quebec university, similar results were reported (Perusse-Lachance et al., 2010). A dietary intake and physical activity study conducted among students (n=134) enrolled in an undergraduate nutrition course in Alberta found that a high percentage of students met guidelines for physical activity (83%) but were not meeting the Dietary Reference Intake (DRI) for vitamin D, vitamin E, and potassium and two-thirds of participants had an excessive intake of sodium (Frehlich et al., 2017). Seemingly, the measures imposed to curb the COVID-19 pandemic can provide an impetus for poor dietary intake and sedentary activity among students. The current COVID-19 pandemic presents a unique opportunity to examine its impact on students’ lives. A healthy diet and regular physical activity are important determinants of health; therefore, understanding whether these modifiable behaviours should be targeted as points of intervention for improving health during future pandemics is critical. The purpose of this study is to examine the impact of COVID-19 on university students’ dietary intake, physical activity and sedentary behaviour. We hypothesized that the COVID-19 pandemic would increase students’ sedentary behaviours, reduce level of physical activity, and negatively impact dietary intake.

## Materials and Methods

### Design and Participants

A survey design consisting of a questionnaire administered pre-COVID-19 (retrospective) and then again during the COVID-19 pandemic (mainly prospective, but retrospective for students recruited later during the pandemic – see description below) was used to examine university students’ dietary intake, physical activity, and sedentary behaviour before and during COVID-19. To collect retrospective data before COVID-19, participants were instructed to complete the ‘before COVID-19 questionnaire’ and consider at least a month (for diet) and a week (for physical activity and sedentary behaviour) before the pandemic in responding to questions. Similarly, to collect prospective data, participants were instructed to complete the ‘during COVID-19 questionnaire’ and consider at a typical month (for diet) and a week (for physical activity and sedentary behaviour) during the pandemic, ensuring a period characterized by strict measures (i.e., April) was captured. Participants were recruited from two universities within the Province of Saskatchewan, Canada: University of Saskatchewan and University of Regina.

The survey targeted students who were enrolled (fulltime) before September 2019, living on their own or with their partner or roommates and responsible for buying and preparing their own meals. Students were excluded from the study if they were new to university (i.e., started university in September 2019 or later), living with parents or guardian, receiving regular prepared meals from any source or if their accommodation included a meal plan. The inclusion and exclusion criteria were indicated on the recruitment emails. Potential participants who qualified for the study were invited to contact a research team member (LB) to obtain an identification number. The recruitment email also contained two different hyperlinks to the questionnaires; one to collect retrospective data of dietary intake, physical activity, and sedentary behaviour before the COVID-19 pandemic and the other to collect prospective data during the pandemic. The recruitment email, inviting students to voluntarily participate, was sent to all colleges and student associations at the University of Saskatchewan (U of S) and faculties at the University of Regina (U of R). Additionally, posters were also advertised through social media and the U of S website portal for announcements. The study was approved by the Biomedical Research Ethics Board of the Universities of Saskatchewan and Regina. The first page of the questionnaire contained an information sheet outlining the study protocols for data collection and maintaining confidentially as well as the consent process. By completing and submitting the questionnaire, participants conveyed that they understood the conditions of participation and their consent was implied. Participation in the study qualified students to enter a draw to win one of 40 gift cards valued at $25 CDN.

### Questionnaire design and measures

The before and during COVID-19 questionnaires included three sections: sociodemographic, physical activity and sedentary behavior, and dietary intake.

#### Sociodemographic

variables included sex, undergraduate or graduate student, year in program, citizenship (Canadian, permanent resident status, international student), living arrangements (live alone, with roommates, or with spouse), transportation (own car or public transport), employment, and source of food.

#### Dietary intake

was assessed using items from the Canadian Diet History Questionnaire II (CDHQII) (National Institutes of Health, 2020). This 165-item past-month food frequency questionnaire of food, beverages and dietary supplements was adapted for use among Canadian populations by Csizmadi et al. (2007; 2016). The CDHQII has been validated for online use among Canadians at a population level (Horne et al., 2020). In assessing the validity of the CDHQII, Horne et al. (2020) demonstrated that there were no significant differences between the dietary intake from the CDHQII and dietary intake from 24-hour recalls (p<0.05). Data were collected on the frequency of consumption and usual portion size of food, beverages, and dietary supplements consumed over a month. Frequency of consumption was captured as how often the food, beverages and dietary supplements were consumed per month, week or day or never. The data collected were entered into the Food Processor® Nutrition Analysis software (ESHA Research, Version 11.1. Salem, Oregon, USA) to estimate the participants’ daily energy, macronutrient, and micronutrient intake, as well as prevalence of inadequacy.

#### Physical activity

was assessed using items from the Godin Leisure-Time Exercise Questionnaire (GLTEQ) (Godin and Shepard, 1985), which measures the number of times during a 7-day period a participant engages in strenuous (vigorous), moderate and mild exercise. The GLTEQ is validated and reliable for use among adult populations (Godin and Shepard, 1985; Miller and Freedson, 1994). The GLTEQ has been validated against other objective measures of physical activity such as the Caltrac accelerometer (r = 0.45, p<0.01) (Miller and Freedson, 1994). Test-retest reliability coefficients for the GLTEQ measures were 0.48, 0.46, 0.94 for light, moderate and strenuous exercise respectively (Godin and Shepard, 1985). The reported scores for strenuous and moderate exercise were used to categorize participants who were meeting the Canadian 24-Hour Movement Guidelines for Adults aged 18–64 years of 150 minutes of moderate to vigorous physical activity per week (Ross et al., 2020).

#### Sedentary activity

was measured by asking participants to report the “average number of hours spent engaging in sedentary activities daily before/during COVID-19”. Sedentary activity was defined as activities spent in a sitting or reclining position while awake such as sitting in class (face to face and online), watching TV, using phone, using the computer, doing assignments, reading, doing crafts or hobbies, sitting around with friends and/or family, or playing music. The reported daily sedentary time was used to categorize participants who were meeting the Canadian 24-Hour Movement Guidelines for Adults of ≤ 8 hours of sedentary time per day (Ross et al., 2020).

### Procedures

The questionnaires were administered online through SurveyMonkey® platform (San Mateo, California, USA). A convenience sample of five students was used to pilot-test the questionnaires. As no changes were made to the items of the questionnaire, the data obtained from these five students were included in the analysis. All participants were instructed to first complete the ‘before COVID-19 questionnaire’, then allow a minimum break of three days before completing the ‘during COVID-19 questionnaire’. Where necessary, reminders to complete both questionnaires were sent 5 days and 7 days after each participant completed the first survey. Additionally, a final reminder was sent to non-respondents of the second questionnaire approximately one-week before closing the survey. Data were collected from April to July 2020. However, participants who enrolled in the study in June and July were instructed to refer to the month of April or May when completing the ‘during COVID-19 questionnaire’ as there were stricter mitigation measures in Saskatchewan at that time.

### Statistical Analysis

Chi-square tests were conducted to determine whether there were differences in sociodemographic variables between those who completed both questionnaires and those who did not. Data (dietary intake, physical activity and sedentary behaviour) were analyzed only for those participants who completed both questionnaires. Descriptive statistics were used to report sociodemographic characteristics of those who completed both questionnaires. Differences in the mean daily energy, macronutrient, and micronutrient intake, physical activity and sedentary activity were analyzed using a sex (male vs. female) x time (before COVID-19 vs. during COVID-19) analysis of variance with repeated measures on time. Bonferroni post-hoc tests were used to assess differences between means when there was a sex x time interaction. The prevalence of inadequacy of vitamins and minerals with an Estimated Average Requirement (EAR) was estimated using the EAR cut-point method. This method determines the proportion of the sample with usual nutrient intakes below the EAR (IOM, 2000). Statistical significance was set at p<0.05. All statistical analyses were completed using Statistica 5.0 (StatSoft, Tulsa, Oklahoma).

## Results

Of the 158 participants who indicated an interest in participating in the survey, 125 participants completed both questionnaires (i.e., participation rate of 79%). There were no differences in the sociodemographic variables between those who completed both questionnaires and those who did not. The final sample (i.e., those who completed both questionnaires) was predominantly composed of students from the University of Saskatchewan (92%). While 55.2 % of the final sample were employed before the COVID-19 pandemic, 48.8% were employed during the pandemic. Additional sociodemographic data of the sample that completed both questionnaires are presented in Table 1.

**Table 1.** Participant sociodemographic characteristics

| Characteristic |  | No. (%) |
| --- | --- | --- |
| Sex | Female | 95 (76) |
|  | Male | 30 (24) |
| Level of Education | Undergraduate | 64 (51.2) |
|  | Post-Graduate | 61 (48.8) |
| Resident Status | Canadian | 88 (70.4) |
|  | Permanent resident | 4 (3.2) |
|  | International student | 33(26.4) |
| Living Arrangement | Live alone | 29 (23.2) |
|  | Live with roommates | 38(30.4) |
|  | Live with partner and/or family | 54 (43.2) |
|  | Live in university's residence | 4 (3.2) |
| Primary Transportation | Own a vehicle | 79 (63.2) |
|  | Public transportation | 33 (26.4) |
|  | Other | 13 (10.4) |

### Dietary Intake

Table 2 represents daily mean frequency of consumption of food before and during COVID-19, by food group. The frequency of consumptions decreased across all food groups during COVID-19 (p<0.05).

**Table 2.** Daily mean frequency of consumption of food before and during COVID-19, by food group.

| Food group | Before COVID-19 (times per day) | During COVID-19 (times per day) |
| --- | --- | --- |
| Beverages | 1.26 | 0.67 |
| Coffee and Tea | 3.32 | 2.02 |
| Grains | 1.03 | 0.92 |
| Fruits | 1.01 | 0.86 |
| Vegetables | 1.37 | 0.75 |
| Dairy | 1.77 | 0.98 |
| Nuts | 0.84 | 0.33 |
| Meat | 1.53 | 1.21 |
| Meat alternatives | 0.68 | 0.39 |
| Snacks | 1.18 | 0.81 |

Table 3. shows the self-reported mean daily energy, macro- and micronutrient, alcohol, and caffeine intake of university students before and during COVID-19. There were significant time main effects for most dietary variables, with the mean intake of most nutrients decreasing during COVID-19, except for alcohol, which increased. No statistically significant differences were found in the mean intake of iodine (p=0.23), and vitamin C (p=0.66) during COVID-19. Also, no significant sex main effects were found reflecting that dietary intake were not significantly different between males and females. There was a significant interaction between the effects of sex and time on the mean intake of cholesterol, vitamin B3 and zinc. Bonferroni post hoc analyses showed that decreases in the intake of zinc (p<0.001), vitamin B3 (p<0.001), and cholesterol (p=0.01) during COVID-19 was significantly greater in males than females. Figure 1 presents the prevalence of inadequacy of vitamins and minerals with an EAR. The prevalence of inadequacy increased for all nutrients during COVID-19. Several nutrients (vitamin B1, vitamin B2, vitamin B3, vitamin B6, vitamin C, iron, phosphorous, zinc, folate, and selenium) had low (<10%) prevalence of inadequacy before COVID-19. Among these, the prevalence of inadequacy for vitamin B12, vitamin C, and folate increased to more than 10% during COVID-19. The prevalence of inadequate intake for magnesium, zinc and calcium was below 20% before COVID-19; however, the prevalence increased to more than 30% during COVID-19. Over 35% of the sample had inadequate intake of vitamin E and D before and during COVID-19.

**Table 3.**
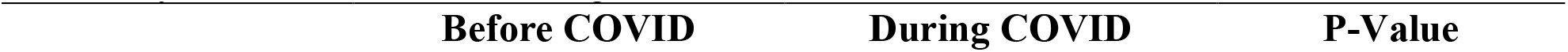
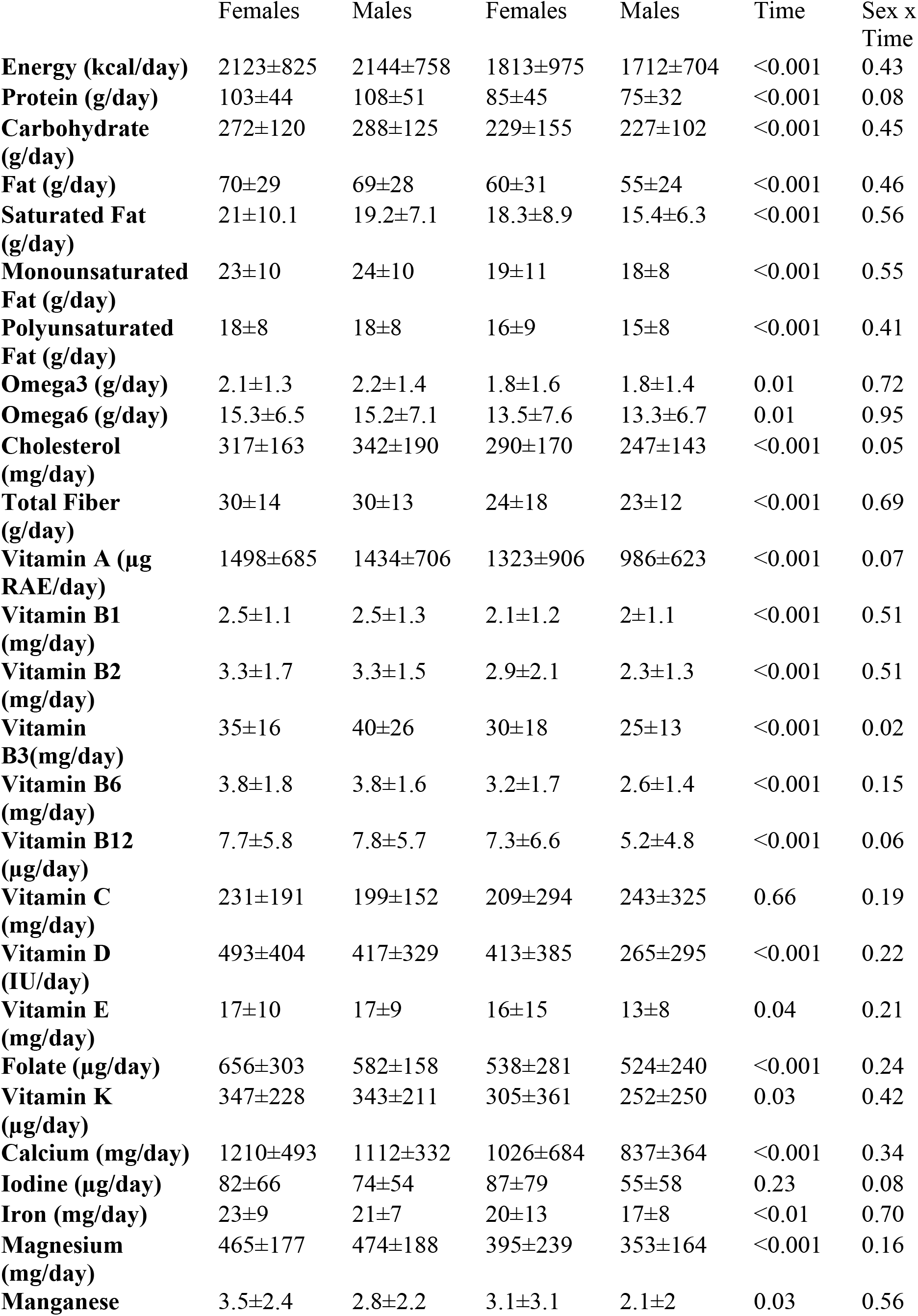
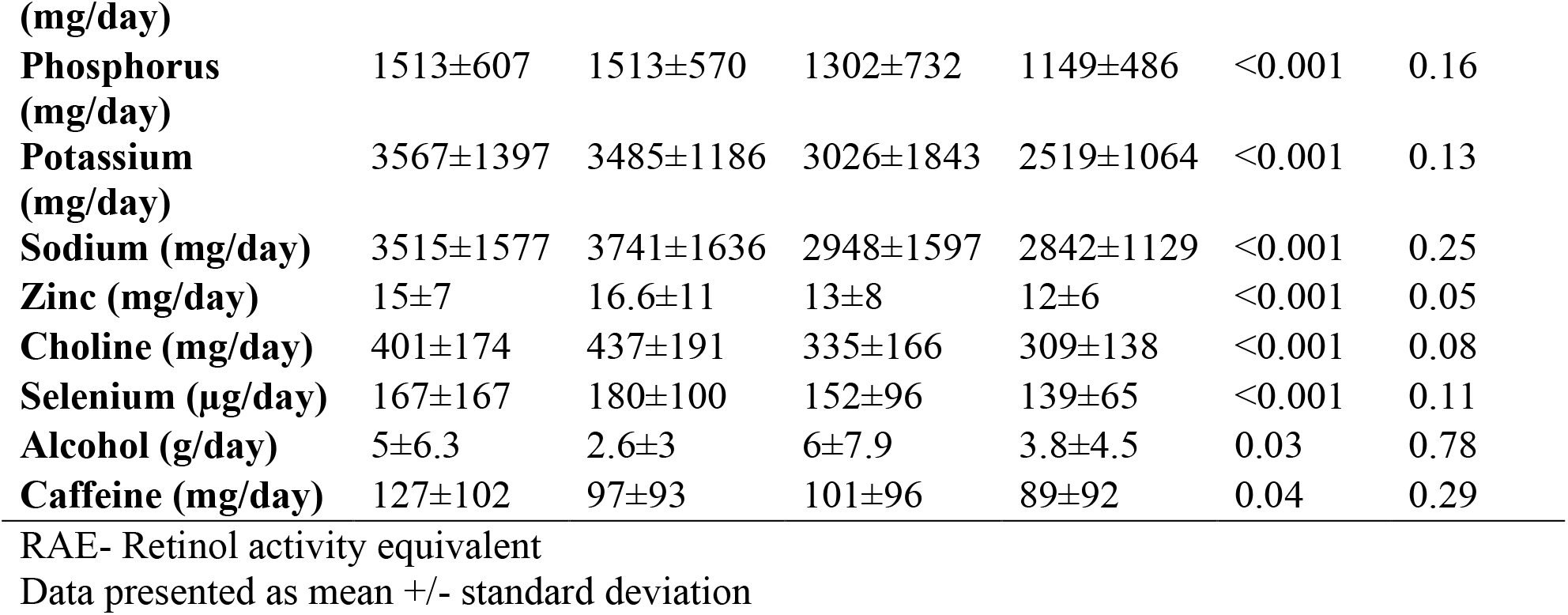
The mean daily energy, macro- and micronutrient, alcohol, and caffeine intake of university students before and during COVID-19

**Figure 1.**
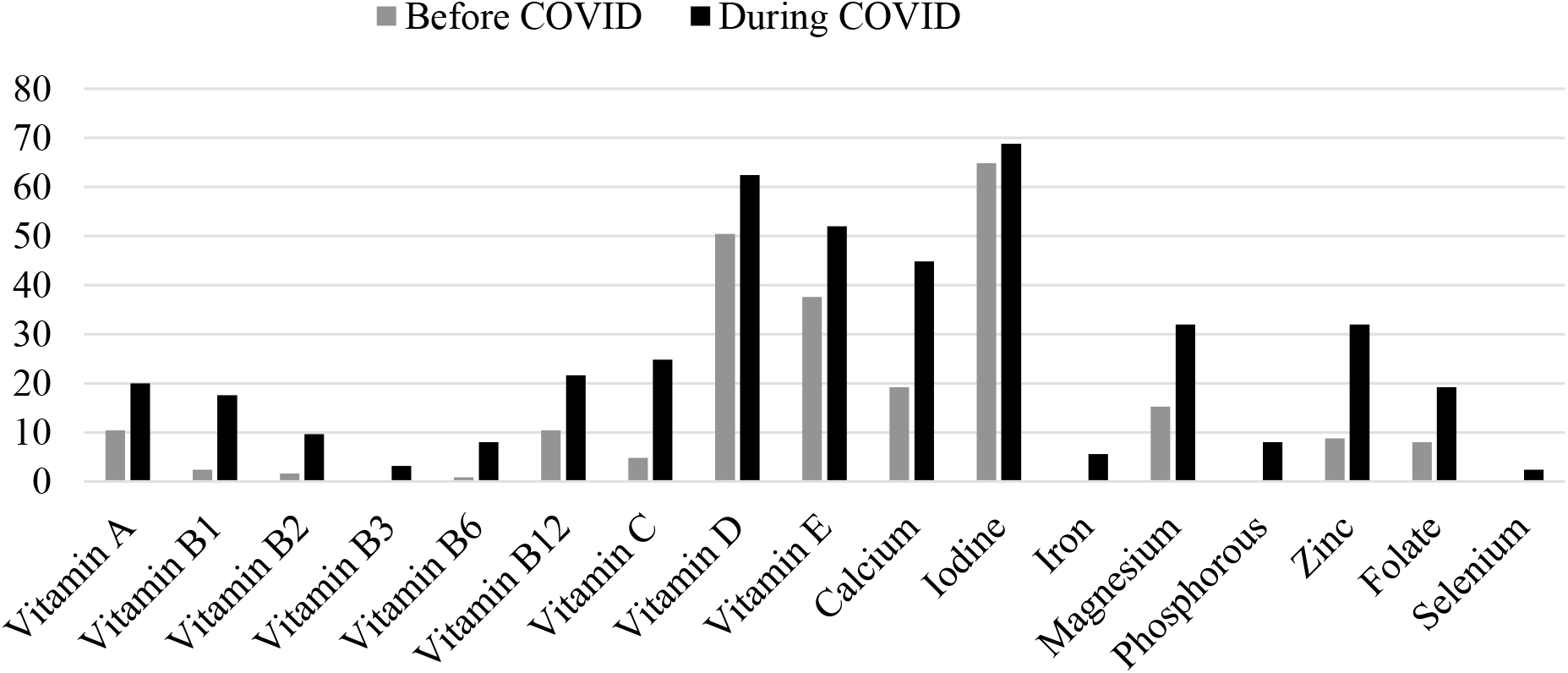
Prevalence of inadequacy of micronutrients with an EAR, before and during the COVID-19 pandemic (%). EAR-Estimated Average Requirement

### Physical and Sedentary Activity

While 16% of the participants were meeting the Canadian 24-Hour Movement Guidelines for Adults (150 minutes of moderate to intense physical activity per week) before the COVID-19 pandemic, only 9.6% met the guidelines during the pandemic. Of the participants who were meeting the guidelines before the pandemic, 90% became less active during the pandemic while 10% became more active. On the other hand, of the participants who were not sufficiently active (i.e., not meeting the guidelines), 55% of them became less active while 45% became more active. Also, the minutes per week spent engaging in moderate to vigorous intensity physical activity decreased from 85±58 minutes to 65±53 minutes (p<0.001). A significant sex main effect was observed for the minutes per week spent engaging in moderate to vigorous intensity physical activity (p=0.01). Compared to their male counterparts females spent more time engaging in moderate to vigorous intensity physical activity. The duration of moderate to vigorous intensity physical activity decreased from 89.4±70 minutes per week to 73.3±55.3 minutes per week among females and from 72.5±62.5 minutes per week to 40.5±35.7 minutes per week among males. The mean number of hours spent in sedentary activities increased from 8.3±3 hours per day before COVID-19 to 11±4 hours per day during COVID-19 (p<0.001). While 54% of the participants were meeting the Canadian 24-Hour Movement Guidelines for Adults’ sedentary behaviour (≤ 8 hours per day) before the COVID-19 pandemic, only 30% met the guidelines during the pandemic.

## Discussion

This study examined the impact of COVID-19 on university students’ dietary intake, physical activity and sedentary behaviour. The findings provide evidence to support our hypotheses, i.e., the COVID-19 pandemic significantly increased students’ sedentary behaviour, reduced their level of physical activity, and negatively affected their nutrient and caloric intake.

Overall, the quality of students’ diet was poorer during COVID-19 given the decreased frequency of consumption of grains, fruits, vegetables, dairy, nuts, meat and meat alternatives. The decreased consumption of these nutrient-dense foods explains the decrease in macro- and micronutrient intake and the increase in the prevalence of nutrient inadequacy observed during COVID-19. The apparent decrease in overall caloric intake can also explain the changes in nutrient intake observed in our study. In contrast to our findings, caloric intake increased among female university students in Australia as a result of increased snacking behaviour during confinement (Gallo et al., 2020). Ammar et al. (2020) also found that during confinement students’ consumption pattern shifted towards more frequent snacking and uncontrolled eating. While other studies reported increased snacking and decreased alcohol consumption during confinement (Ammar et al., 2020; Gallo et al., 2020), our study found that there was significantly less frequent snacking while alcohol consumption increased significantly. Along with decreased consumption of nutrient dense foods and increased snacking, increased alcohol consumption contributes to poor diet quality.

A diet rich in essential micronutrients is beneficial to psychological health (Collins et al., 2020; Lassale et al., 2019), immune function (Calder, 2020) and overall health. Moreover, during adverse events like the current COVID-19 pandemic, good nutritional quality can reduce susceptibility to disease (Calder, 2020). Unfortunately, our findings show a significant decrease in the intake of most macro- and micronutrients during confinement. What is more concerning is that many of the essential vitamins and minerals show a high prevalence of inadequacy both before and during confinement. For example, vitamin D had an alarmingly high prevalence of inadequacy before and during COVID-19 despite the heightened interest in this nutrient’s role in reducing the severity of the disease (Mattioli et al., 2020; Ebadi and Montano-Loza, 2020). Vitamin C also has a significant role to play in immune function (Lee, 2019; Carr and Maggini, 2017); however, despite it being one of the few nutrients with no statistically significant change in intake during COVID-19, there was a significant increase in the prevalence of inadequate intake of vitamin C during COVID-19. Our study showed that during COVID-19, the prevalence of inadequate intake increased for several other nutrients (calcium, magnesium, zinc) important for bone health (Price et al., 2012). On a public health level, many Canadian adults (19 years and older) have inadequate intakes of these micronutrients (Health Canada, 2012). This increased inadequate micronutrient density combined with long hours of sedentary behaviour and inadequate physical activity poses a three-fold risk to bone health during COVID-19.

Several possibilities can explain the dietary shift observed during confinement. Experiences of psychological distress, as is possible during a pandemic, have been consistently linked to poor diet quality, particularly increased consumption of alcohol, decreased consumption of nutrient-dense foods, or increased consumption of high fat and sugar foods (Mattioli et al., 2020; Tomiyama, 2019; Quehl et al., 2017; Macht, 2008; Torres & Nowson, 2007). Also, reducing caloric intake may be one mechanism through which our study participants balanced their energy during COVID-19. Considering that physical activity levels decreased, and sedentary activity increased, participants expended less energy. Likely too, is that provincial measures implemented to prevent the spread of the COVID-19 virus impacted students’ dietary intake by means of their food access and/or availability. Once the provincial State of Emergency was declared in response to the COVID-19 pandemic, both universities transitioned to remote teaching while in the wider public grocery stores reduced their hours of operation and capacity for shoppers (Government of Saskatchewan. 2020). The resulting home confinement, closures of businesses, and reduced hours of operation of food stores, may have limited students’ shopping frequency and at-home food availability as well as reduced their exposure and access to community and organizational food environments.

Despite recommendations from the Canadian 24-Hour Movement Guidelines for Adults aged 18–64 years to engage in at least 150 minutes of moderate to vigorous physical activity per week to achieve health benefits (Ross et al., 2020), only 16% of our study participants were meeting this recommendation before COVID-19. This value fits well with an earlier report on the proportion of Canadians (15.4%) meeting the physical activity guidelines (Colley et al., 2011). However, during COVID-19, only 9.6% of our participants were meeting this recommendation. Given that the months of April and May, 2020, marked a period when the province of Saskatchewan enforced very strict measures in response to the COVID-19 pandemic, there is no doubt that measures such as the closures of gyms and other recreational facilities by the universities and other private and public establishments within the province (Government of Saskatchewan, 2020) resulted in reductions in the level of physical activity. Other possible explanations for the reduction in physical activity may be that many students were no longer actively commuting to school given that the Universities of Saskatchewan and Regina transitioned to remote learning (Government of Saskatchewan, 2020). Compared to their adult counterparts in our study, a greater proportion of Canadian children (5-11 years) and youth (12-17), 23.8% and 13.2% respectively, were meeting their age specific guidelines for physical activity during COVID-19 (Moore et al., 2020). Other studies also provided evidence to support the negative impact that COVID-19 has on physical activity. In one multi-continent study, the frequency, duration and intensity of physical activity decreased by 35%, 34% and 42.7% respectively during home confinement (Ammar et al., 2020). Lesser and Nienhuis (2020) conducted a survey among Canadian adults and found that 40.5% of inactive participants in their study became less active, while 33% became more active. A similar finding was also observed in our study whereby 90% of the participants who were meeting the guidelines before the pandemic became less active during the pandemic. These findings imply that interventions geared at increasing physical activity during a pandemic should target active as well as inactive individuals, and also ensure that both groups engage in a level of physical activity that aligns with guidelines for health benefits. There is much potential to increase physical activity during a pandemic as our study also demonstrated that 45% of the participants who were not sufficiently active (i.e., not meeting the guidelines) became more active. It is possible that such persons increased their level of physical activity to offset the increases in sedentary behaviour. The observed increase in physical activity in some participants could also be explained by easier access to exercise equipment at home or greater support to become more active during home confinement. With regards to the sex differences in physical activity, our findings contradict previous reports in the literature indicating that males are more active than females (Colley et al., 2011; Trost et al., 2002). In fact, we found that females had greater physical activity levels before and during COVID-19. The reason for this contradictory finding is not entirely clear but could be interpreted as being a result of a disproportionate ratio of females to males in our sample. Women accounted for 76% of our sample. Another plausible explanation could be that the motivation (peer pressure, weight loss or maintenance) for physical activity among females led to higher levels of physical activity particularly during university enrollment when body weight is likely to increase.

Consistent with expectations, COVID-19 brought an increase in the average number of hours (8.3 to 11 hours) students spent engaging in sedentary activities. It is also remarkable that over half (54%) of the sample were meeting the Canadian 24-Hour Movement Guidelines for Adults’ sedentary time (i.e., ≤ 8 hours) before the pandemic but not during the pandemic. Although modest, the statistically significant increase in sedentary behaviour agrees with findings from other studies which investigated the impact of COVID-19 on adults (Ammar et al., 2020) and children (Moore et al., 2020). The observed changes in sedentary behaviour points to the likelihood that the switch in the delivery of university classes from on-campus to remote during COVID-19 (Government of Saskatchewan, 2020) resulted in students spending more time sitting. Sedentary time would also be affected by the public health recommendation in Saskatchewan (Government of Saskatchewan, 2020) to ‘stay home’. The average sedentary time during COVID-19 reported by participants in our study is consistent with the duration of sedentary time reported among university students in Ontario, Canada in a pre-pandemic condition (Prapavessis et al., 2015). Moreover, the increase in sedentary time is not surprising given that the COVID-19 pandemic favors many of the factors that promote sedentary behaviours among university students, including physical fatigue, state of mind, intentions to be sedentary, social modelling, peer pressure, residency, computer and TV access, moved class attendance, exams, and assignments (Deliens et al., 2015; Prapavessis et al., 2015). Of particular concern is that not only did participants’ sedentary time shift from being approximate to that which is recommended (∼ 8 hours) within the Canadian 24-Hour Movement Guidelines for Adults (Ross et al., 2020) before the pandemic, but the average duration of sedentary time (11 hours) reported during the pandemic has been associated with a variety of unfavorable health outcomes including metabolic syndrome (Edwardson et al., 2012), reduced cognitive function (Falck et al., 2017) diabetes, cardiovascular disease, and all-cause mortality (Ekelund et al., 2019; Biwas et al., 2015, Wilmot et al., 2012). Ekelund et al. (2016) suggest that sedentary behaviour and physical activity are related although disparate, and that regular moderate intensity physical activity (60–75 min per day) is beneficial in mitigating the health risks associated with prolonged sitting. Therefore, despite being in home confinement during COVID-19; students can engage in regular moderate intensity physical activity to help mitigate the health risks associated with prolonged sitting; however, our findings suggest that during COVID-19 students did not engage in sufficient physical activity to offset the risks associated with the increased sedentary behaviour brought about by home confinement.

To our knowledge this is the first study to assess the impact of the COVID-19 pandemic on dietary intake, physical activity and sedentary behaviour concurrently on the same sample. In interpreting the findings of this study, several limitations need to be considered. The data describing dietary intake, physical activity and sedentary behaviour during COVID-19 represent a period of time when there were stricter mitigation measures in Saskatchewan, Canada; thus, the data should be interpreted in the context of this period. Pre-COVID-19 data were collected retrospectively, thus participants were required to recall information related to their dietary intake, physical activity levels and sedentary behaviour prior to the period of data collection. As a result, recall bias may have led to an underestimation or overestimation in the data presented this study. Additionally, considering that the pre-COVID-19 recall period may have led participants to recall their health behaviour during the winter season, seasonal variation in these health behaviours should also be considered. The data presented in this study were self-reported by participants, therefore, consistent with this approach, social desirability may have affected the results. Despite the fact that recruitment at both campuses was campus-wide and a large number of students was theoretically reached, 125 completed the survey. As a result, the findings of this study may not be reflective of the greater student population at the Universities of Saskatchewan and Regina. The number of participants may be due to the possibility that students had less interest in participating given the shift in the mode of delivery to remote learning and the associated challenges faced dealing with the adjustments to the COVID-19 pandemic.

## Conclusion

Our findings confirm that university students’ poor dietary intake, low physical activity, and high sedentary behaviour were already risks for poor health before the pandemic. These risks were significantly compounded during the pandemic. University students should be targeted for interventions aimed at maintaining and improving physical activity and dietary practices during this pandemic and beyond.

## Data Availability

All data is available from the authors upon request.

## Data availability statement

All data is available from the authors upon request.

